# Evaluating the PATHFAST TB LAM Ag Assay as a Treatment Monitoring Tool for Pulmonary Tuberculosis : Protocol for a Prospective Longitudinal Study in Nairobi, Kenya

**DOI:** 10.1101/2025.11.07.25339800

**Authors:** Yu Takaizumi, Joy Kinoti, Mayu Hikone, Fred Orina, Hellen Meme, Jane Ong’ang’o, Betty Muriithi, Elizabeth Mueni, Satoshi Kaneko, Emily Lai-Ho MacLean, Shuntaro Sato, Nobuo Saito

**Author notes:** These authors contributed equally to this work (Yu Takaizumi, Joy Kinoti, Mayu Hikone). Corresponding author Nobuo Saito.

## Abstract

**Background:** Treatment failure remains a major challenge in tuberculosis (TB) management. Rapid and objective assessment of treatment response is essential, as existing tools have limited accuracy and slow turnaround times. The PATHFAST TB LAM Ag assay (PATHFAST-LAM), an automated chemiluminescent enzyme immunoassay, was developed to quantify lipoarabinomannan (LAM) in sputum within one hour. Previous studies have shown a strong correlation between sputum LAM concentration and culture-based bacterial load. However, its clinical utility for predicting poor outcomes during treatment has not been prospectively evaluated.

**Methods and analysis:** We will conduct a prospective longitudinal study enrolling newly diagnosed, bacteriologically confirmed pulmonary TB patients at Rhodes Chest Clinic and Mbagathi County Referral Hospital in Nairobi, Kenya. We will follow participants throughout the 6-month treatment course, attempting to collect sputum weekly during weeks 1–4, biweekly during weeks 5–12, and monthly during months 3–6. We will measure LAM concentrations at these time points using the PATHFAST-LAM assay. The primary outcome is to assess whether changes in sputum LAM concentration during the intensive phase (baseline to week 4 and/or week 8) predict a composite poor outcome, defined as positive sputum culture at month 6, treatment failure, death during treatment, or relapse within three months after treatment completion. The primary endpoint is the area under the curve (AUC) from the receiver operating characteristic (ROC) analysis, representing the predictive performance of changes in sputum LAM concentration for the composite poor outcome. We will identify the optimal cut-off value for LAM change and estimate sensitivity and specificity with 95% confidence intervals using 2×2 tables. We will apply an adaptive design that allows sample-size re-estimation after interim analysis.

**Ethics and dissemination:** The study was approved by the Kenya Medical Research Institute (KEMRI/SERU/CRDR/124/5241) and Nagasaki University (250619327), and registered at ClinicalTrials.gov (NCT07157904). Findings will be disseminated through peer-reviewed publications and scientific meetings.

**Article Summary:** *Strengths and limitations of this study:*

- PATHFAST TB LAM Ag Assay (PATHFAST-LAM) automatically measures LAM in sputum and provides rapid, quantitative results within an hour, requiring minimal manual handling.
- This is the first prospective longitudinal study to evaluate sputum LAM concentrations as measured by PATHFAST-LAM in relation to clinical outcomes among patients with pulmonary tuberculosis (PTB).
- The study’s prospective design allows real-time assessment of sputum LAM dynamics throughout treatment and follow-up, providing valuable data on temporal changes linked to treatment outcomes.
- This study enables clinical evaluation of the potential of PATHFAST-LAM as a new treatment monitoring tool (TMT) for PTB.
- The cost-effectiveness and feasibility of implementation in routine practice are not fully evaluated in this study.

## Introduction

Tuberculosis (TB) continues to pose a major global health burden, affecting millions of people each year despite being a curable disease (1). Effective treatment regimens are available, but the long duration of therapy and difficulties in maintaining adherence often lead to treatment failure (1). As treatment failure results not only in unfavorable outcomes for patients but also in the emergence and transmission of drug-resistant TB, early and accurate assessment of treatment response is crucial (2,3). The World Health Organization (WHO) recommends sputum culture and acid-fast bacilli (AFB) smear microscopy to monitor treatment response in pulmonary TB (PTB) (4). However, culture has a long turnaround time and requires advanced laboratory facilities, while AFB smear microscopy has limited sensitivity and cannot distinguish viable from non-viable bacilli (5,6). To address these limitations, the WHO Target Product Profile (TPP) for TB treatment monitoring tools (TMTs) highlights the need for advanced tools that are accurate, rapid, and feasible for use in resource-limited laboratory settings (7).

Lipoarabinomannan (LAM), a glycolipid component of the *Mycobacterium tuberculosis* (MTB) cell wall, has mainly been evaluated as a rapid urine-based diagnostic test (8,9). However, its application as a TMT has not yet been established. Recent studies have explored the potential of LAM concentration in sputum as a biomarker for the TMT. In a study of PTB patients, Kawasaki et al. reported that sputum LAM concentrations measured by enzyme-linked immunosorbent assay (ELISA) correlated with bacterial load determined by the Mycobacteria Growth Indicator Tube (MGIT) culture system (10). To simplify the complex ELISA procedure, the PATHFAST TB LAM Ag assay (PATHFAST-LAM; PHC Corporation, Tokyo, Japan) has been developed as a fully automated, cartridge-based chemiluminescent enzyme immunoassay (CLEIA) that quantifies sputum LAM within an hour. The study showed that sputum LAM concentrations measured by PATHFAST-LAM correlated with bacterial load determined by the MGIT system (11). Two studies from South Africa and Kenya also found similar results (12,13). Furthermore, these studies demonstrated treatment-related declines in sputum LAM measured by PATHFAST-LAM. The study in Kenya showed that the decreases were particularly evident among patients with high baseline levels (12), while the study in South Africa found that sputum LAM declined in parallel with bacterial load determined by culture-based methods during the first 14 days of treatment (13). However, these findings were based on retrospective analyses using stored samples, and the clinical utility of PATHFAST-LAM for detecting treatment failure has not been evaluated prospectively. The aim of this study is to prospectively assess whether sputum LAM concentrations measured by PATHFAST-LAM are associated with treatment outcomes in patients with PTB.

## Methods and analysis

### Design

This is a prospective longitudinal study conducted to determine whether changes in sputum LAM concentrations during tuberculosis treatment can predict poor treatment outcomes in Kenya.

### Setting

We will conduct this study in Nairobi, Kenya, at two major healthcare facilities that serve as key centers for tuberculosis care. The study sites are Rhodes Chest Clinic (RCC) and Mbagathi County Hospital (MCH). RCC is a specialized outpatient clinic dedicated to tuberculosis management and treats approximately 700–800 new PTB cases each year. MCH is a large tertiary-level public hospital that provides both outpatient and inpatient tuberculosis services, managing about 1,000 new TB cases annually with a bed capacity of around 150. If we encounter an insufficient number of cases or an inadequate overall sample size, we will recruit additional participants from other healthcare facilities in Nairobi.

### Participants

We will include adult inpatients and outpatients aged 18 years or older with newly diagnosed PTB confirmed by sputum Xpert MTB/RIF Ultra, who have not received any TB treatment within the previous six months. Participants must be able to provide written informed consent, attend scheduled follow-up visits, and be willing to attempt to provide sputum samples.

### Recruitment and enrollment

Healthcare workers at the study sites will screen adults who present symptoms suggestive of PTB and have positive results from sputum Xpert MTB/RIF Ultra and/or AFB smear microscopy. After confirming eligibility, our research staff will explain the study procedures and obtain written informed consent from participants or their legal representatives before initiating any study-related activities. If Xpert MTB/RIF Ultra testing has not been performed as part of routine care, our research team will perform the test after provisional enrollment and exclude participants with negative Xpert results.

### Outcome measures

We define a composite poor outcome as any of the following: (1) positive sputum culture at month 6, (2) treatment failure, (3) death during treatment, or (4) relapse within three months after treatment completion (Table 1) (3).

**Table 1.** Definition of composite poor outcome in this study

| Outcome | Definition |
| --- | --- |
| Composite poor outcome | Patients who meet any of the following criteria:<br>• Death from any cause<br>• Discontinuation of TB treatment for $\geq 2$ consecutive months<br>• Early relapse (within 3 months after completing TB treatment)<br>• Change in TB treatment regimen for any reason<br>• No culture conversion at the end of TB treatment |

### Data collection and follow-up

We will collect clinical, microbiological, and outcome data during treatment and follow-up as outlined in Figure 1 and Supplementary Table 1.

**Figure 1.**
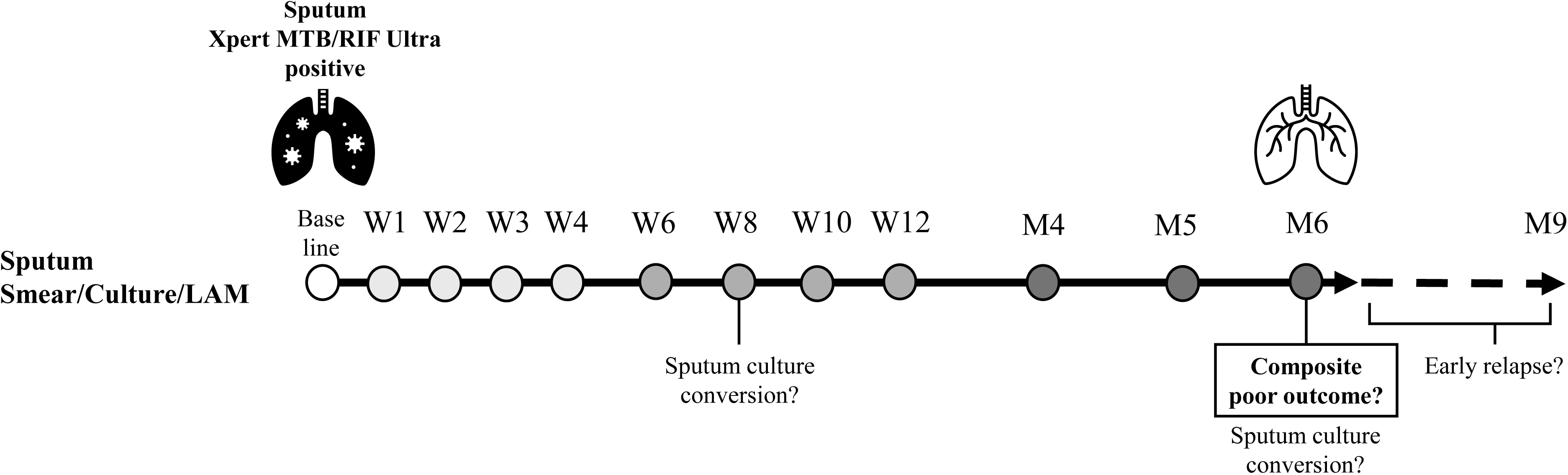
Schedule of sample collection and outcome assessment in this study. Sputum for LAM measurement was collected weekly (W1–4), biweekly (W5–12), and monthly (M3–6). Treatment outcomes were evaluated at treatment completion. W, week; M, month.

### Baseline assessment

At enrollment, we will obtain demographic and clinical information using a structured questionnaire and review of medical records. We will collect 5 mL of sputum as baseline samples. Sputum samples will undergo AFB smear microscopy, MGIT culture, and drug susceptibility testing (DST) using Xpert MTB/RIF Ultra and MGIT DST.

### Follow-up visits

We will follow participants weekly during weeks 1–4, biweekly during weeks 5–12, and monthly during months 3–6 of treatment (Figure 1). At each visit, we will collect 5 mL of sputum. Sputum testing will include AFB smear microscopy, culture, and LAM measurement. We will continue sputum collection until spontaneous production ceases. After treatment completion, we will follow up with participants by telephone monthly for three months to check for early relapse. If early relapse is suspected, we will ask them to return to the clinic for evaluation and sample collection.

### Outcome assessment

We will perform clinical evaluations at months 1, 2, 3, and 6 during treatment, and will follow up with participants by telephone monthly for three months after treatment completion. Based on clinical, microbiological, and radiological findings, we will assess treatment outcomes according to the National Tuberculosis, Leprosy and Lung Disease Program (NTLD-P) definitions (14). We will perform whole-genome sequencing on baseline and relapse isolates to differentiate relapse from reinfection.

### Laboratory analysis

We will measure LAM concentrations in sputum using the PATHFAST-LAM assay (PHC Corporation, Tokyo, Japan) following the manufacturer’s instructions and previously validated procedures (11). Each sample will be analyzed on the day of collection or the following day. We will process both raw sputum and (N–acetyl–L–cysteine–sodium hydroxide (NALC–NaOH) processed sputum samples. Previous studies have shown no significant difference in LAM concentrations between raw and NALC–NaOH processed sputum, although supporting data are limited (13). Because laboratories that perform culture testing often process sputum using the NALC–NaOH whereas front-line TB clinics use raw sputum for smear microscopy or Xpert MTB/RIF testing, we will measure both types to evaluate the assay’s applicability across different laboratory settings. For each test, we will mix 200 µL of sputum with 100 µL of 1 N NaOH, heat the mixture at 100°C for 20 minutes, neutralize it with 50 µL of 5 M NaHDPOD, and centrifuge it at 3,000 × g for 5 minutes at 25°C. We will collect the supernatant as the LAM extract. The extraction process requires approximately 30 minutes. We will transfer 100 µL of the LAM extract into the reagent cartridge and load it into the PATHFAST-LAM analyzer, which automatically performs the measurement. The analytical principle and detailed procedures have been described elsewhere (11,13).

### Sample size calculation

As no previous studies have evaluated the association between sputum LAM reduction and treatment outcomes in PTB, a formal sample size calculation based on prior effect estimates is not feasible. Therefore, we will adopt an adaptive sample size approach, incorporating an initial feasibility exploration phase followed by sample size re-estimation based on interim analysis. We plan to enroll an initial cohort of 300 participants in a year, based on operational feasibility and presumptive sample size calculation (15). Calculated sample size indicated that 49 patients with composite poor outcomes are required. Given that 20% of PTB patients will experience poor outcomes in our setting (3), 245 patients would be required. Allowing for a 15–20% loss to follow-up, the estimated sample size for the feasibility cohort is 300 participants. An interim analysis will be performed when the initial 30 participants have completed the outcome measures at the 6-month follow-up. The sputum LAM reduction at key time points (baseline to 4 weeks and 8 weeks) will be calculated. Based on the calculated LAM reduction, the distribution and variability of sputum LAM reduction will be analyzed to explore appropriate cut-off values for classifying treatment response. Then, sensitivity will be calculated based on varied cutoff values, and the target sensitivity will be determined. Based on the interim analysis, the total sample size will be determined to achieve a one-sided width of ±10% for the 95% CI, using Wilson’s score method based on the target sensitivity.

### Statistical analysis

As no established cutoff value exists for changes in sputum LAM concentration to predict a composite poor outcome, we will determine an optimal threshold through receiver operating characteristic (ROC) analysis.

The composite outcome at 6 months will be analyzed as a binary variable (poor outcome vs. favorable outcome). Sensitivity and specificity will be calculated across different cutoff values of LAM change, and the optimal cutoff will be identified. Based on this cutoff, we will classify participants as test-positive or test-negative for predicting the composite poor outcomes. We will then construct a 2×2 contingency table and calculate sensitivity, specificity, and the corresponding 95% confidence intervals to quantify the predictive performance of the assay.

### Ethics and dissemination

The study protocol and informed consent documents have been approved by KEMRI, Scientific and Ethics Review Unit (SERU) (KEMRI/SERU/CRDR/124/5241) and Institute of Tropical Medicine, Nagasaki University (250619327). Written informed consent will be obtained from all participants. Confidentiality will be ensured through anonymized data.

## Discussion

### Significance and expected outcomes

This prospective longitudinal study conducted in Nairobi, Kenya aims to evaluate sputum LAM measured by the PATHFAST-LAM assay as a TMT for PTB. By following patients longitudinally through treatment and post-treatment phases, the study seeks to determine whether early changes in sputum LAM concentration can predict poor treatment outcomes. If validated, PATHFAST-LAM could provide a rapid, quantitative, and accessible means for treatment monitoring, enabling timely clinical interventions and reducing the risk of treatment failure. If the assay shows good predictive performance, future studies should explore its potential role in clinical management and treatment decision-making.

### Strengths of the study

A major strength of this study is its prospective and longitudinal design, allowing real-time assessment of sputum LAM dynamics throughout treatment and follow-up. It is the first to evaluate whether changes in sputum LAM concentrations can predict clinical treatment outcomes, extending beyond previous studies that examined only microbiological correlations (11–13). Conducting this study in Kenya, a high-burden setting for TB and HIV, under real-world clinical conditions by trained research staff, will provide valuable evidence on the feasibility and utility of PATHFAST-LAM in resource-limited settings. The study may include patients with varying disease severity and recruits from different healthcare levels, enhancing its generalizability and clinical relevance. All participants will undergo sputum culture and drug susceptibility testing, enabling detection of drug-resistant TB and assessment of its association with LAM dynamics.

### Limitations

This study has several limitations. First, the proportion of multidrug-resistant or extensively drug-resistant TB cases in Kenya is relatively low (7.1% in 2023), and the number of treatment failures due to multi drug resistance in this study may therefore be limited (16). However, poor adherence and other factors can also contribute to treatment failure, and the expected rate of approximately 20% should provide sufficient cases for analysis (12,16). Second, although we will follow up participants by telephone for three months after treatment completion to monitor for early relapse, poor outcomes occurring beyond this period will not be captured because follow-up will not be extended further. Third, the cost-effectiveness and feasibility of routine implementation are difficult to evaluate, as PATHFAST testing is performed by the research team rather than facility staff, and feasibility in resource-limited or rural settings is also difficult to assess. Lastly, LAM measurement may not be feasible in patients who are unable to produce sputum, and sputum collection may become difficult after clinical improvement during treatment.

## Author Statement

### Author’s contribution

YT, JK, MH, BM, LM, SS, and NS conceived the study and developed the study design. YT, MH, and NS drafted the study protocol and wrote the initial manuscript. FO, HM, JO, BM, EM, and NS critically reviewed and revised the manuscript for important intellectual content. FO, HM, JO, and EM provided clinical expertise and contributed to the adaptation of study procedures to the local setting. NS supervised the overall study. All authors read and approved the final version of the manuscript. The study protocol was written by YT, JK, MH, BM and NS. YT, MH and NS wrote the initial manuscript. FO, HM, JO, BM, SK, EM and NS critically reviewed the draft of the manuscript. All authors have read and approved the final version of the manuscript.

### Funding statement

The work is supported by Japan Agency for Medical Research and Development (AMED) Grant Number (24jk0110031h0001) to Nobuo Saito.

### Competing interests

PHC Corporation (Tokyo, Japan) will provide PATHFAST TB LAM Ag assay kits free of charge and will rent the PATHFAST analyzer to the research team. PHC Corporation will not be involved in the study design, data collection, analysis, interpretation, manuscript preparation, or decision to publish. No other financial support or incentives will be received.

### Data availability statement

All data produced in the present study are available upon reasonable request to the authors

## Notes

### Competing Interest Statement

The authors have declared no competing interest.

### Clinical Trial

NCT07157904

### Clinical Protocols

https://clinicaltrials.gov/study/NCT07157904

### Author Declarations

The study protocol and informed consent documents have been approved by the Kenya Medical Research Institute (KEMRI), Scientific and Ethics Review Unit (SERU) (KEMRI/SERU/CRDR/124/5241) and Institute of Tropical Medicine, Nagasaki University (250619327).

