## Supplementary Table 1 for "Evaluating the PATHFAST TB LAM Ag Assay as a Treatment Monitoring Tool for Pulmonary Tuberculosis : Protocol for a Prospective Longitudinal Study in Nairobi, Kenya"

**Supplementary Table 1. Schedule of study activities in this study**

| Activity | **Base**  **line** | **W1-3** | **W4** | **W6** | **W8** | **W10** | **W12** | **M4** | **M5** | **M6** |
| --- | --- | --- | --- | --- | --- | --- | --- | --- | --- | --- |
| **Clinical information** | | | | | | | | | | |
| Demographic information | 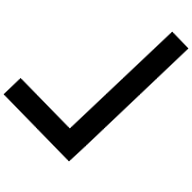 |  |  |  |  |  |  |  |  |  |
| Clinical interview  TB symptoms | 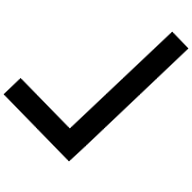 |  | 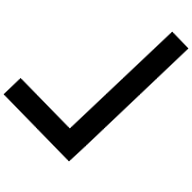 |  | 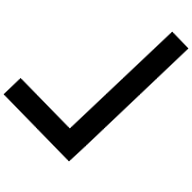 |  | 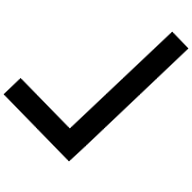 |  |  | 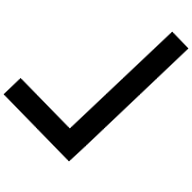 |
| Physical examination  Vital signs / Body weight | 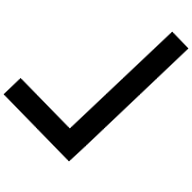 |  | 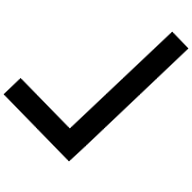 |  | 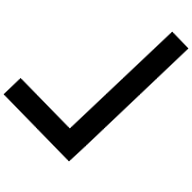 |  | 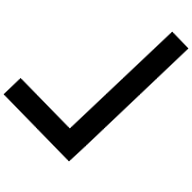 |  |  | 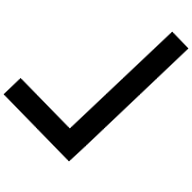 |
| **Laboratory** | | | | | | | | | | |
| Sputum  Xpert MTB/RIF Ultra | 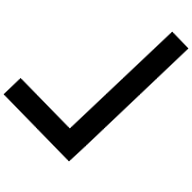 |  |  |  |  |  |  |  |  |  |
| Sputum AFB/culture | 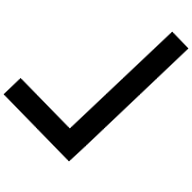 | 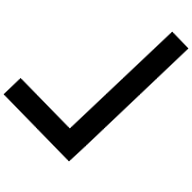 | 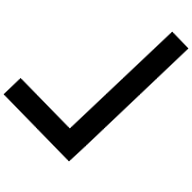 | 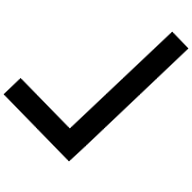 | 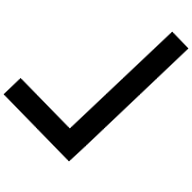 | 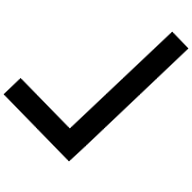 | 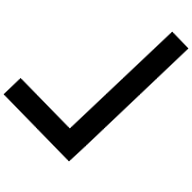 | 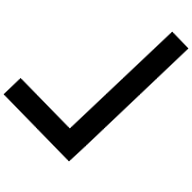 | 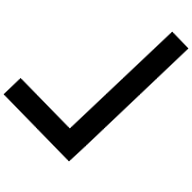 | 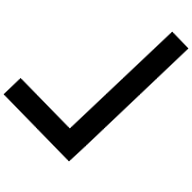 |
| LAM (sputum) | 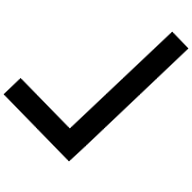 | 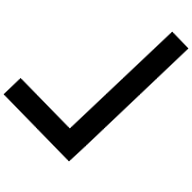 | 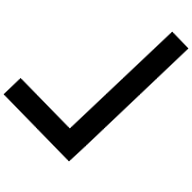 | 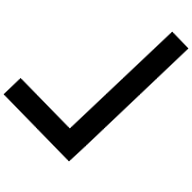 | 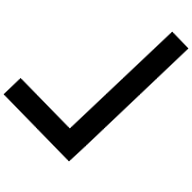 | 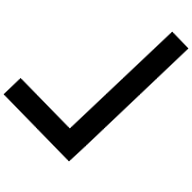 | 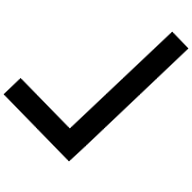 | 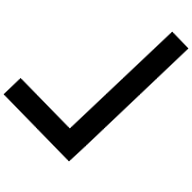 | 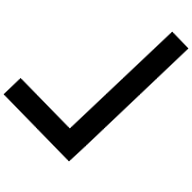 | 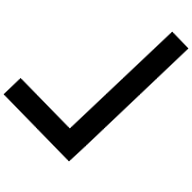 |
| **Outcomes** | | | | | | | | | | |
| Composite poor outcome |  |  |  |  |  |  |  |  |  | 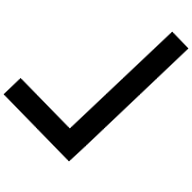 |
| TB programmatic outcome |  |  |  |  |  |  |  |  |  | 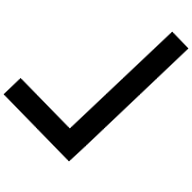 |
| Sputum culture conversion |  |  |  |  | 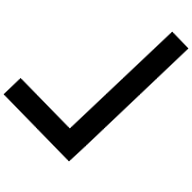 |  |  |  |  | 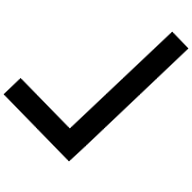 |

W, week; M, month; AFB, acid-fast bacilli; LAM, lipoarabinomannan; TB, tuberculosis.
“Clinical interview” includes assessment of TB symptoms. “Composite poor outcome” is defined in Table 1 of the main text.
